# Group A Streptococcus Molecular Point of Care testing in a Paediatric Emergency Department

**DOI:** 10.64898/2026.04.20.26351279

**Authors:** Ewurabena A. Mills, Rebekah Bingham, Ruud G. Nijman, Shiranee Sriskandan

**Affiliations:** Department of Infectious Disease, Faculty of Medicine, Imperial College London, London, UK; Department of Paediatric Emergency Medicine, Division of Medicine, St. Mary’s Hospital, Imperial College NHS Healthcare Trust Diseases, Imperial College London, UK; Centre for Paediatrics and Child Health, Imperial College London, UK; NIHR Health Protection Research Unit in Healthcare Associated Infections and AMR, Imperial College London, London, UK; Centre for Bacterial Resistance Biology, Imperial College London, London, UK

**Author notes:** Corresponding author Shiranee Sriskandan, Dept. of Infectious Diseases, Hammersmith Hospital Campus, Du Cane Road, London W12 0NN, UK.

## Abstract

**Background:** An upsurge in *Streptococcus pyogenes* infections 2022-2023 highlighted potential benefits of point-of-care tests (POCT) to support clinical pathways, prevent outbreaks, and optimise antibiotic use.

**Objectives:** We conducted a pilot research study in a west London paediatric emergency department (ED) to determine whether a molecular POCT had potential to alter management in children who were also having a conventional throat swab taken for culture.

**Methods:** Children <16 years presenting to ED who had a throat swab requested by a clinician were invited to have a second swab taken for research purposes only. Clinical management was unaffected by the research swab result, which was processed using a molecular POCT that was not approved for use in the host NHS Trust.

**Results:** Prevalence of streptococcal infection was low during the study (May 2023-June 2025); swab positivity in symptomatic children was 12.8% (6/47). Overall, 38/49 (77.6%) participants who had throat swabs received antibiotics. Of those children recommended to receive antibiotics, 29/38 (76.3%) had a negative POCT. Mean time to reporting of positive throat swab culture results was 3.67 days (range 3-5 days) leading to occasional delay in treatment, although POCT identified positive results within minutes.

**Conclusion:** Antibiotic use was frequent and could be avoided or stopped by use of a ‘rule out’ POCT in over three-quarters of children in the ED, if suspicion of *S. pyogenes* is the main driver for prescribing. POCT were easy to process and produced immediate results compared with culture, in theory enabling timely decision-making and avoiding treatment delay.

## Introduction

In England and Wales, national guidance advocates against diagnostic testing for sore throat, instead recommending use of syndromic scoring systems, such as FeverPain or Centor score, to identify patients over 5 years who might benefit from antibiotics (1). NICE guidance in 2008 advocated a pragmatic no-treatment or deferred treatment approach to sore throat based on syndromic assessment only (2). Point of care testing (POCT) for *Streptococcus pyogenes* (group A *Streptococcus*, GAS) is undertaken in many other developed countries in Europe and North America to aid diagnosis (3,4). However, predicted minimal effects on reducing antibiotic use have precluded adoption in England and Wales in those aged over 5 years (5), with no specific evaluation in younger children, where scoring systems are of uncertain value (6).

A post-pandemic upsurge of *S. pyogenes* in 2022–2023 renewed interest in POCT. In England, there were 58,972 scarlet fever notifications, 3,729 invasive GAS infections, and 491 deaths, including 55 in children (7), alongside increased beta-lactam prescribing (8) and supply chain pressures. Interim guidance (9) lowered thresholds for antibiotic use and allowed alternatives under a serious shortage protocol. Public concern as well as disease surge drove major service pressures, with ED attendances rising sharply for sore throat (+953%) and scarlet fever (+2640%) compared with winter means (10).

To investigate whether the rapid molecular detection of *S. pyogenes* might have potential to optimise ED management of children presenting with sore throat, we undertook a research and feasibility study in west London. Children with suspected infection who had a throat swab taken for culture were invited to have a second swab for research purposes.

## Methods

### Recruitment

The study ran from 25^th^ May 2023 to 25^th^ June 2025. Children aged <16 years of age presenting to the paediatric ED at Imperial College Healthcare NHS Trust with any acute febrile syndrome, and where the attending clinician requested a throat swab for culture were invited to take part in the study. Parents or guardians were approached by the research team to ask if a second (research) swab could be taken. The research was approved by a National Research Ethics Committee (Camden and Kings Cross 14/LO/2217 IRAS 166513).

The original aim was to recruit 200 children during the period when routine guidelines were interrupted; however NICE guidelines were re-instated in February 2023, and swabbing practice consequently reduced. The study was therefore terminated after 50 children were recruited for pragmatic reasons.

### Microbiology and other data collected

Routine throat swabs were collected at the clinician’s request and transported in Amies medium to the hospital laboratory for standard culture and reporting. *S. pyogenes* was confirmed using MALDI-TOF (Bruker Biotyper). Research swabs from consenting participants were tested separately by a researcher using a molecular POCT (Abbott ID Now). As this test lacked local NHS Trust validation, results were not shared with clinicians and were for research only.

Data collected included demographics, presentation time, ED stay duration, FeverPAIN and Centor scores, prescriptions, and outcomes (e.g. admission or representation). Culture results, including time to result and subsequent prescribing actions, were also recorded.

## Results and Discussion

Data from 49 children who had both clinical and POCT swabs taken were included. Clinical swab culture results on two children were missing. The average age was 5.8 years (range 9 months-15 years).

Several observations emerged. Antibiotic prescribing was common (38/49, 77.5%) among children who had a throat swab, likely reflecting concern for bacterial infection (Table 1). Clinicians suspected *S. pyogenes* across a range of presentations in the ED; indications for throat swabbing included sore throat (with or without fever, or with lymphadenitis) (n=24); rash with or without fever or cough (n=11); fever only (n=9); other (n=5, including lymphadenitis and cough). Cough is considered a negative predictor of *S. pyogenes* in current sore throat scoring algorithms, but may need re-evaluation given the risks of pneumonia/empyema in younger children (7). In the group studied, scoring systems appeared to have been of limited value, including those with sore throat (Supplementary Table 1) albeit these children were swabbed perhaps because clinical scoring was deemed unreliable.

**Table 1.** Antibiotic decision for whole cohort in the ED.

|  | Received antibiotic | Did not receive antibiotic* |
| --- | --- | --- |
| Number (% of total) | 38 (77.6) | 11 (22.4) |
| Already on antibiotic | 11 | 2 |
| FeverPAIN** Score mean (range) | 2.38 (1-4) | 2.45 (1-3) |
| Centor** Score mean (range) | 1.89 (0-3) | 1.9 (1-3) |
| Throat culture positive | 5 | 1 |
| Time to culture result, mean in days (range) | 3.25 (1 - 8) | 3 (1 - 5) |
| DNA POCT positive | 8 | 2 |
| DNA POCT fail | 1 | 0 |
| DNA POCT negative | 29 | 9 |

\*includes one patient receiving a prescription for delayed antibiotic
\*\*FeverPAIN and Centor scores collate clinical signs and symptoms. NICE guidance advocates antibiotics to be considered for Centor $\geq 3$ (out of 4) or FP $\geq 4$ (out of 5) in people >5 years of age (1).

The overall prevalence of throat swab positivity was 12.8% (6/47) in symptomatic children, consistent with reduced prevalence of streptococcal infection during the study period (May 2023-June 2025). Of the 6 children who had a positive *S. pyogenes* throat culture, 5 of these would have been identified with the DNA POCT immediately, compared with a 3-5 day delay waiting for a culture result (Table 2). In one case, the POCT delivered an indeterminate result due to need for a software upgrade. Of 6 children with positive throat culture results, all bar one received empiric antibiotics; that child had to be recalled 3 days later when the culture result was received, underlining the value of a rapid result.

**Table 2.** Characteristics of whole cohort by throat culture results*.

|  | *S. pyogenes* culture positive | *S. pyogenes* culture negative |
| --- | --- | --- |
| Total | 6 | 41 |
| DNA POCT positive | 5 | 5 |
| DNA POCT negative | 0 | 36 |
| DNA POCT fail | 1 | 0 |
| Time to culture result, mean in days (range) | 3.67 (3-5) | 3.13 (1-7) |
| FeverPAIN** score, mean (range) | 2.2 (0-3) | 2.42 (1-4) |
| Centor** score, mean (range) | 2.4 (2-3) | 1.80 (1-3) |
| Given antibiotics | 5 | 31 |
| Not given antibiotics | 1 | 10 |
| Given analgesics | 1 | 21 |
| Unknown | 1 | 6 |
| Not given analgesics | 4 | 14 |

\*Excludes 2 patients with missing results from throat swab culture
\*\*FeverPAIN and Centor scores collate clinical signs and symptoms. NICE guidance advocates antibiotics to be considered for Centor $\geq 3$ (out of 4) or FP $\geq 4$ (out of 5) in people >5 years of age

Of the 41 children who had a negative throat culture result, 31/41 (75.6%) received antibiotics initially. However, of those who received antibiotics, 27/31 (87.1%) had a negative DNA POCT test, and might have avoided antibiotic treatment altogether had a POCT been used and the result acted upon (Table 2). We cannot exclude the possibility that another type of bacterial infection was suspected in children presenting to the ED, providing an alternate rationale for antibiotic prescribing. Focusing only on the 24 children with sore throat as a presentation (Supplementary Table 2), 19/24 received antibiotics initially, however of these, 14/19 (74%) had a negative POCT result and would have avoided antibiotics. The time taken for a culture result to be available was highly variable, ranging up to 7 days depending on when the swab was taken, meaning that some children remained on antibiotics for up to a week without need.

The POCT was positive in 5 cases where the routine swab for culture was negative. If culture is considered to be the gold standard, then these might be considered ‘false positives’ and trigger unnecessary prescribing. Discordance could be explained by the fact that swabs for culture and POCT were not always taken by the same person nor at exactly the same time. Two children with discordant results were taking antibiotics already. A POCT that detects pathogen DNA will remain positive despite a negative culture result, a feature that could be useful to aid decision making about continuation of antibiotics. The remaining discordant results all occurred in June 2023 at a time when the upsurge in *S. pyogenes* was tailing off, potentially reflecting a higher prevalence of infection. After June 2023, there were no further culture-negative, POCT-positive results. Balanced against the value of a negative POCT in preventing unnecessary prescribing (n=27), the hazards of children with a positive POCT being prescribed unnecessary antibiotics appears minimal, at least in this low prevalence setting.

It is estimated that in primary care, 59% of consultations for sore throat in England result in prescription of an antibiotic (11). ‘Sore throat test and treat’ and ‘Pharmacy First’ studies of pharmacy-led POCT in the community have been conducted in Wales and Northern Ireland respectively (12,13). Both studies evaluated use of POCT in patients aged >5 (>6 in Wales) years with a FeverPAIN score of 2 or more to guide prescribing. Overall, 21%-25% of consultations resulted in an antibiotic prescription, which is much lower than primary care in England, where POCT are not used (11). These studies used antigen detection tests, that are cheaper than molecular tests and many do not require a dedicated instrument. A Cochrane review also found that antibiotic prescribing could be reduced by 25% if POCT were used (14).

A previous implementation study in London used scoring system-directed molecular testing and found that antibiotic use for tonsillitis was less than the previous year (15). Trials outside the UK support the use of POCT in paediatric ED (16). Our study was conducted to determine feasibility of a larger trial of POCT in a UK ED setting. Performing the molecular test was easy for staff to do and results were obtained within minutes. Of 119 children screened for the study, only 49 were recruited mainly due to reluctance to have a second research-only swab. However, parents were keen to have a diagnostic test. We posit that there would be increased willingness to participate if results were to be provided at the time of swabbing to clinicians and patients.

The pilot study intentionally utilised a molecular POCT as it was felt that clinicians would wish to use the most sensitive test to effectively rule out *S. pyogenes* infection, although current cost may preclude broader use. Whether a negative antigen test would provide similar assurance is unclear. Overall, the results overwhelmingly point to a benefit of POCT in order to reduce unnecessary prescribing in the paediatric ED setting at least during our study period. The results also highlight the rapidity of *S. pyogenes* detection compared with conventional culture, a feature that could be of enormous value in optimising time to treatment, controlling outbreaks, and supporting ED pathways. Economic analyses are needed to balance testing costs against value of optimised prescribing, including both reduction of transmission as well as avoidance of unnecessary antibiotic use.

## Data Availability

All data produced in the present study are available upon reasonable request to the authors

## Funding

This study was undertaken as an NIHR Health Protection Unit (HPRU) in Healthcare Associated Infections and AMR response to the upsurge in group A streptococcal infections and received NIHR HPRU funds to purchase the Abbot ID Now instrument and kits. It was also funded by an ICHT Charity Fellowship to EM.

## Acknowledgements

We thank the Paediatric ED research nursing team for their support in patient recruitment. The authors thank the NIHR Biomedical Research Centre (BRC) at Imperial College London that funds the Colebrook laboratory where POCT were initially evaluated *in vitro*.

## Transparency

The authors declare no conflicts of interest.

## Supplementary Tables

**Supplementary Table 1.** Antibiotic decision for children presenting with sore throat.

|  | Received antibiotic | Did not receive antibiotic |
| --- | --- | --- |
| Number (% of total) | 19 (79.2) | 5 (20.8) |
| Already on antibiotic | 7 | 1 |
| FeverPAIN* Score mean (range) | 2.79 (1-4) | 2.6 (1-3) |
| Centor* Score mean (range) | 1.74 (0-3) | 2 (1-3) |
| Throat culture positive | 1 | 1 |
| Time to culture result, mean in days (range) | 3.17 (1-7) | 3.6 (2-5) |
| DNA POCT positive | 4 | 1 |
| DNA POCT fail | 1 | 0 |
| DNA POCT negative | 14 | 4 |

\*FeverPAIN and Centor scores collate clinical signs and symptoms. NICE guidance advocates antibiotics to be considered for Centor $\geq 3$ (out of 4) or FP $\geq 4$ (out of 5) in people >5 years of age

**Supplementary Table 2.**
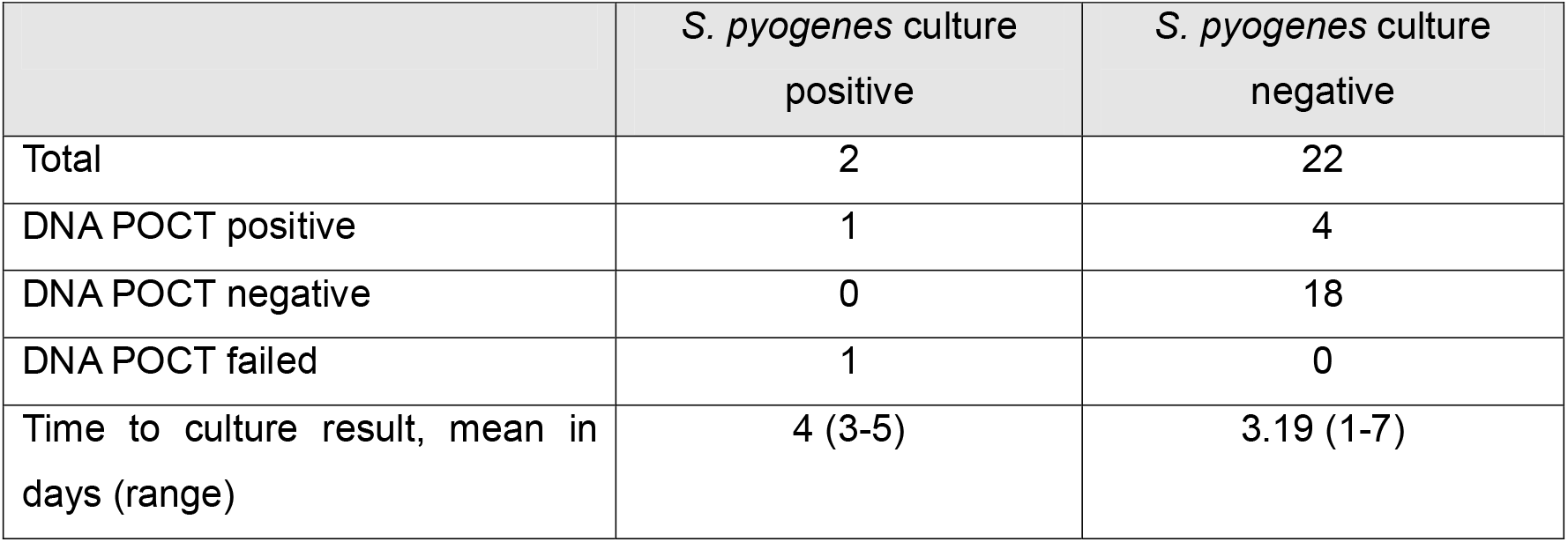
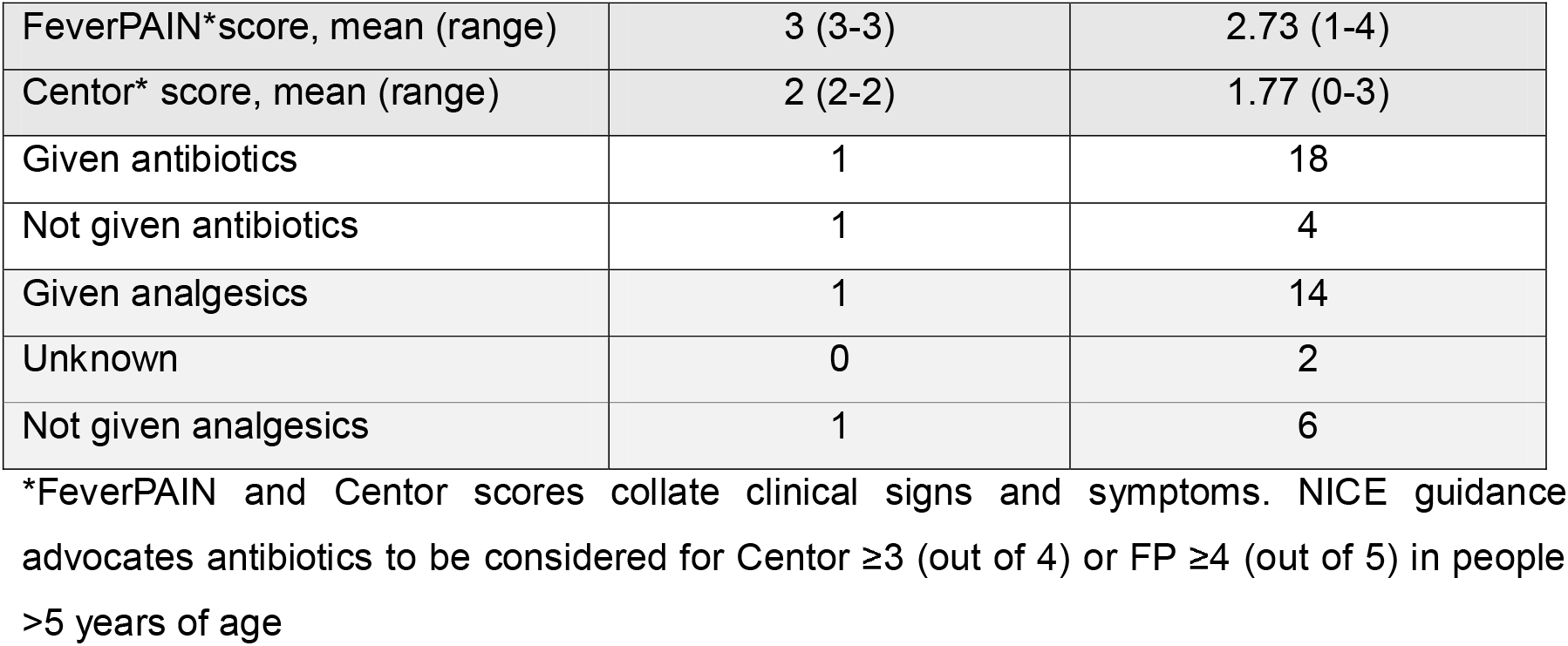
Characteristics of children presenting with sore throat by throat culture results.

## Notes

### Competing Interest Statement

The authors have declared no competing interest.

### Funding Statement

This study received NIHR HPRU funds to purchase the Abbot ID Now instrument and kits. It was also funded by an ICHT Charity Fellowship to EM.

### Author Declarations

The National Research Ethics Committee (Camden and Kings Cross) gave ethical approval for this work (14/LO/2217 IRAS 166513)

